# *UBTF* Tandem Duplications in Pediatric MDS and AML: Implications for Clinical Screening and Diagnosis

**DOI:** 10.1101/2023.11.13.23298320

**Authors:** Juan M. Barajas, Masayuki Umeda, Lisett Contreras, Mahsa Khanlari, Tamara Westover, Michael P. Walsh, Emily Xiong, Chenchen Yang, Brittney Otero, Marc Arribas-Layton, Sherif Abdelhamed, Guangchun Song, Xiaotu Ma, Melvin E. Thomas, Jing Ma, Jeffery M. Klco

## Abstract

Recent genomic studies in adult and pediatric acute myeloid leukemia (AML) demonstrated recurrent in-frame tandem duplications (TD) in exon 13 of upstream binding transcription factor (*UBTF*). These alterations, which account for ~4.3% of AMLs in childhood and up to 3% in adult AMLs under 60, are subtype-defining and associated with poor outcomes. Here, we provide a comprehensive investigation into the clinicopathological features of *UBTF*-TD myeloid neoplasms in childhood, including 89 unique pediatric AML and 6 myelodysplastic syndrome (MDS) cases harboring a tandem duplication in exon 13 of *UBTF*. We demonstrate that *UBTF-*TD myeloid tumors are associated with dysplastic features, low bone marrow blast infiltration, and low white blood cell count. Furthermore, using bulk and single-cell analyses, we confirm that *UBTF*-TD is an early and clonal event associated with a distinct transcriptional profile, whereas the acquisition of *FLT3* or *WT1* mutations is associated with more stem cell-like programs. Lastly, we report rare duplications within exon 9 of *UBTF* that phenocopy exon 13 duplications, expanding the spectrum of *UBTF* alterations in pediatric myeloid tumors. Collectively, we comprehensively characterize pediatric AML and MDS with *UBTF-*TD and highlight key clinical and pathologic features that distinguish this new entity from other molecular subtypes of AML.

**Key Points:**

- Largest cohort of pediatric *UBTF-*TD in myeloid neoplasms reported to date.
- Use of single-cell DNA+protein sequencing technology in 3 *UBTF-*TD samples reveals a clonal evaluation pattern characterized by sequential acquisition of *WT1* and *FLT3* mutations and a more stem cell-like protein expression pattern.
- Pediatric MDS and AML patients with *UBTF-*TD alterations dysplastic features with an increase erythroid precursors.
- Tandem duplications in exon 9 of *UBTF* represent a rare but functionally equivalent subgroup of *UBTF-*TD myeloid neoplasms.

**Impact Statement:** *UBTF* tandem duplications (TD) are subtype-defining genomic alterations in adult and pediatric myeloid neoplasms. Here, we provide a comprehensive characterization of the largest cohort of pediatric *UBTF*-TD cases to date, including the recognition of additional UBTF alterations that mimic the exon 13 duplications in pediatric AML.

## Introduction

Pediatric acute myeloid leukemia (AML) and myelodysplastic syndrome (MDS) are characterized by unique genetic backgrounds when compared to those in adults (1–3). Recurrent tandem duplications (TD) of exon 13 of upstream binding transcription factor (*UBTF*) were only recently identified as potential initiating events in pediatric AML (4–7), accounting for about 4% of newly diagnosed pediatric AML. PCR-based screening covering exon 13 of *UBTF* also identified *UBTF*-TD alterations in large adult AML cohorts (8, 9). These studies significantly contributed to the accumulation of evidence of *UBTF-*TD alterations in adult AML. However, PCR-based methods potentially underestimate partial tandem duplications (PTD) extending outside the regions covered by amplicons or possible alterations not involving exon 13 (9). Also, data on UBTF alterations in pediatric AML is limited to screening of relatively small cohorts (4, 5), and further efforts are needed to accumulate more knowledge about the biology and clinicopathologic features of this disease entity.

*UBTF* encodes the UBTF/UBF protein known to regulate ribosomal RNA (rRNA) transcription and nucleolar formation (10, 11). We previously reported that expression of exon 13 UBTF-TD in cord blood CD34+ (cbCD34+) cells is sufficient to induce cellular proliferation, increase clonogenic activity and the establishment of a transcriptional signature that recapitulates that observed in UBTF-TD AML patient samples (4). Our previous analyses also demonstrated that UBTF-TD do not occur with other canonical alterations in pediatric AML, but that *UBTF*-TD AMLs often harbor additional somatic mutations, such as internal tandem duplications in *FLT3* (*FLT3*-ITD) and frameshift mutations in *WT1*. The acquisition of these cooperating mutations can likely contribute to the stepwise progression of the disease and clonal evolution. However, our understanding of how these cooperating mutations contribute to the cellular and disease status remains to be elucidated.

To bridge these knowledge gaps, we present an extended pediatric cohort of 89 AML and 6 MDS samples with exon 13 *UBTF*-TD, showing that *UBTF-*TD is strongly associated with dysplastic features while acquisition of *FLT3*-ITD is associated with progression from MDS to AML. By leveraging bulk RNA-sequencing and single cell genomics, we show that the co-occurrence of *FLT3*-ITD and *WT1* mutations is associated with stem cell-like phenotypes. Furthermore, we identified tandem duplications within exon 9 of *UBTF* in two cases transcriptionally resembling exon 13 *UBTF*-TD AML. Exon 9 UBTF-TDs contain hydrophobic leucine-rich sequences and can induce leukemic phenotypes in cbCD34+ cells, which recapitulate findings in exon 13 UBTF-TD, suggesting that they likely have a shared mechanism and should be classified as the same molecular entity. These findings offer valuable insights to inform future diagnostic strategies and understanding of the molecular basis of *UBTF*-TD myeloid neoplasms.

## Materials/Subjects and Methods

### Cell Culture and Analysis of cord-blood CD34+ Cell Models

UBTF-TD cbCD34+ models were generated as previously described (4). Freshly isolated cord blood CD34+ (cbCD34+) cells were transduced with lentiviral particles from MND-PGK-mCherry constructs expressing N-terminus HA-tagged UBTF-wild-type (WT), N-terminus HA-tagged UBTF-TD, N-Terminus HA-tagged UBTF-TD78-exon9 or N-Terminus HA-tagged UBTF-TD153-exon9. Transduced cells were sorted for mCherry positivity and expanded as previously described(4). Molecular experiments were performed at 40-60 days after sorting unless otherwise noted. For cytospins, 100,000 cells were washed with 1X PBS and spun onto Superfrost Plus Microscope slides (12-660-16, Fisher Scientific) at 800 rpm for 5 min.

### DNA Single-Cell Sequencing and Analysis

Single-cell targeted sequencing was performed using Tapestri System from Mission Bio (missionbio.com). A panel of 162 PCR amplicons with an average size of 260bp was designed using Tapestri Designer from Mission Bio (designer.missionbio.com), as well as a manually designed amplicon targeting the *UBTF* exon13 TD region. This panel consists of amplicons targeting *UBTF* tandem duplicated regions, common mutations in *UBTF-*TD AML, and mutations specific to cases presented in this manuscript (**Supplemental Table 2**). Cryopreserved UBTF-TD primary AML samples were thawed and subjected to dead cell removal using the EasySep^TM^ Dead Cell Removal Kit (STEMCELL Technologies, cat# 17899). Live cells were then subjected to the Mission Bio DNA+Protein protocol per manufacturer’s instructions (missionbio.com). Libraries were then sequenced on the Novaseq platform (100M read pairs for DNA libraries and 225M read pairs for protein libraries). Bam files, loom files, h5 files, and QC metrics were produced via a customization of the Tapestri pipeline developed by Mission Bio (support.missionbio.com/hc/en-us). Analysis of the samples was completed using the Mosaic package v3.0.1 (missionbio.github.io/mosaic/). Reads for the *UBTF* and *FLT3*-ITD calls were isolated from bam files using the pysam python package v0.21.0 (github.com/pysam-developers/pysam). Reads were then realigned to ITD contigs reported in previous studies (4) using the BWA aligner v0.7.15-r1140 (12, 13). When necessary, mutation VAFs were adjusted using pysam.

### RNA-sequencing and genomic profiling

As we previously performed (1), RNA reads from newly sequenced samples and from publications were mapped to the GRCh37/hg19 human genome assembly using the StrongARM pipeline (14). Chimeric fusion detection was carried out using CICERO (v0.3.0)(15). For somatic mutations calling from RNA-seq BAM files, we applied Bambino (v1.07)(16) for SNV and RNAindel (v3.0.4) (17, 18) focusing on 87 predefined genes recurrently mutated in pediatric AML and myelodysplastic syndrome. UBTF-TD screening was performed as we have reported (4). For the cases with TWIST capture sequencing, we called mutations as previously described (4). We also collected mutation calls with DNA data from referenced publication in Supplemental Table 5.

### Transcriptome analysis

Gene expression analysis was performed as previously described (1). Briefly, an RNA-seq cohort was established by integrating UBTF cases with RNA-seq data in this study (n=96) and AML in other categories from the published study (1) (n=837) and cbCD34+ cells (n=5). Reads from aligned RNA-Seq BAM files were assigned to genes and counted using HTSeq (v0.11.2) (19) with the GENCODE human release 19 gene annotation. The count data were transformed to log2-counts per million (log2CPM) using Voom available from R package Limma (v3.50.3) (20). The top variable genes were selected using the “vst” method in Seurat package (21). The expression data were then scaled, and PCA (Principal Component Analysis) was performed on the scaled data using the top 265 variable genes. Dimension reduction was performed using UMAP (Uniform Manifold Approximation and Projection) (22) with the top 100 principal components. Differential gene expression analysis was performed by Limma between groups as indicated in each figure, and we set Log2 CPM = −1 if it is < −1 based on the Log2 CPM data distribution. *P* values were adjusted by the Benjamini-Hochberg method to calculate the false discovery rate (FDR) using R function p.adjust. Genes with absolute fold change > 2 and FDR < 0.05 were regarded as significantly differentially expressed. Gene Set Enrichment Analysis (GSEA) was performed by GSEA (v4.2.3) using MSigDB gene sets c2.all (v7.5.1) (23). Permutations were done 1000 times among gene sets with sizes between 15 and 1500 genes.

### Statistics

Details about statistical comparisons are provided in each figure legend. All the computations were done using R or GraphPad Prism, and all *P* values are 2-sided.

## Results

### UBTF-TD in pediatric myeloid neoplasms

Our previous study described the molecular features of 27 pediatric AML cases with tandem duplications in exon 13 of *UBTF* (*UBFT*-TD) (4). To expand the cohort and better understand the biology, we screened RNA-sequencing data of pediatric MDS and AML cases and identified an additional 68 cases from available datasets and previously published studies (2, 24) and routine clinical service at St. Jude Children’s Research Hospital. All 95 cases (median age = 14 yrs., range = 2.4-27.4) possessed exon 13 *UBTF*-TDs encoding a consensus hydrophobic leucine-rich ELTRLLARM motif within the duplications (**Figure 1A, Supplemental Table 1**). The duplications resulted in an increased size of exon 13 (median size = 60 bp, range = 45-339) (**Figure 1B**). Consistent with previous findings, *UBTF*-TD did not co-occur with other subtype-defining alterations and showed high variant allele frequencies (VAFs, median = 36.3%, Interquartile range (IQR) = 10.2%), further supporting our previous assertion that *UBTF*-TD alterations are early clonal events (1, 4) (**Figure 1C**). We further investigated the mutational background of this *UBTF*-TD cohort, confirming a strong association with a normal or trisomy 8 karyotype (**Figure 1C-E**), as well as with internal tandem duplications (ITD) in *FLT3* (n=55, 57.9%) and mutations in *WT1* (n=39, 41.1%), which are highly co-occurring (**Figure 1F**, *P*=0.011, Fisher’s exact test) with 30.1% (n=29) cases harboring both alterations. In addition, 26.6% (n=25) of cases also had at least one mutation in Ras-MAPK pathway genes: *NRAS* (n=17,17.9%); *PTPN11* (n=5, 5.3%); *RIT1* (n=5, 5.3%); *NF1* (n=4, 4.2%); *CBL* (n=2, 2.1%); and *KRAS* (n=2, 2.1%). Other recurrent mutations in myeloid malignancies were rarely observed, including *IDH1*/*IDH2* (n=3, 3.2%), *BCOR* (n=2, 2.1%), and *RUNX1* (n=1, 1.1%). Although our previous studies only evaluated AML samples, we also identified *UBTF*-TD in 6 cases of MDS with normal karyotype. These include 3 out of 46 primary MDS cases (6.2%) from our previously published cohort (2). Five of the six MDS cases were classified as childhood MDS with increased blasts according to the current WHO classification (25) and lacked known germline predispositions to pediatric MDS or monosomy 7/del7q. No *FLT3*-ITD mutations were detected in the 6 MDS cases, while *WT1* mutations were present in 3 out of the 6 MDS cases, suggesting clonal evolutionary patterns initiating with *UBTF-*TD alterations.

**Figure 1.**
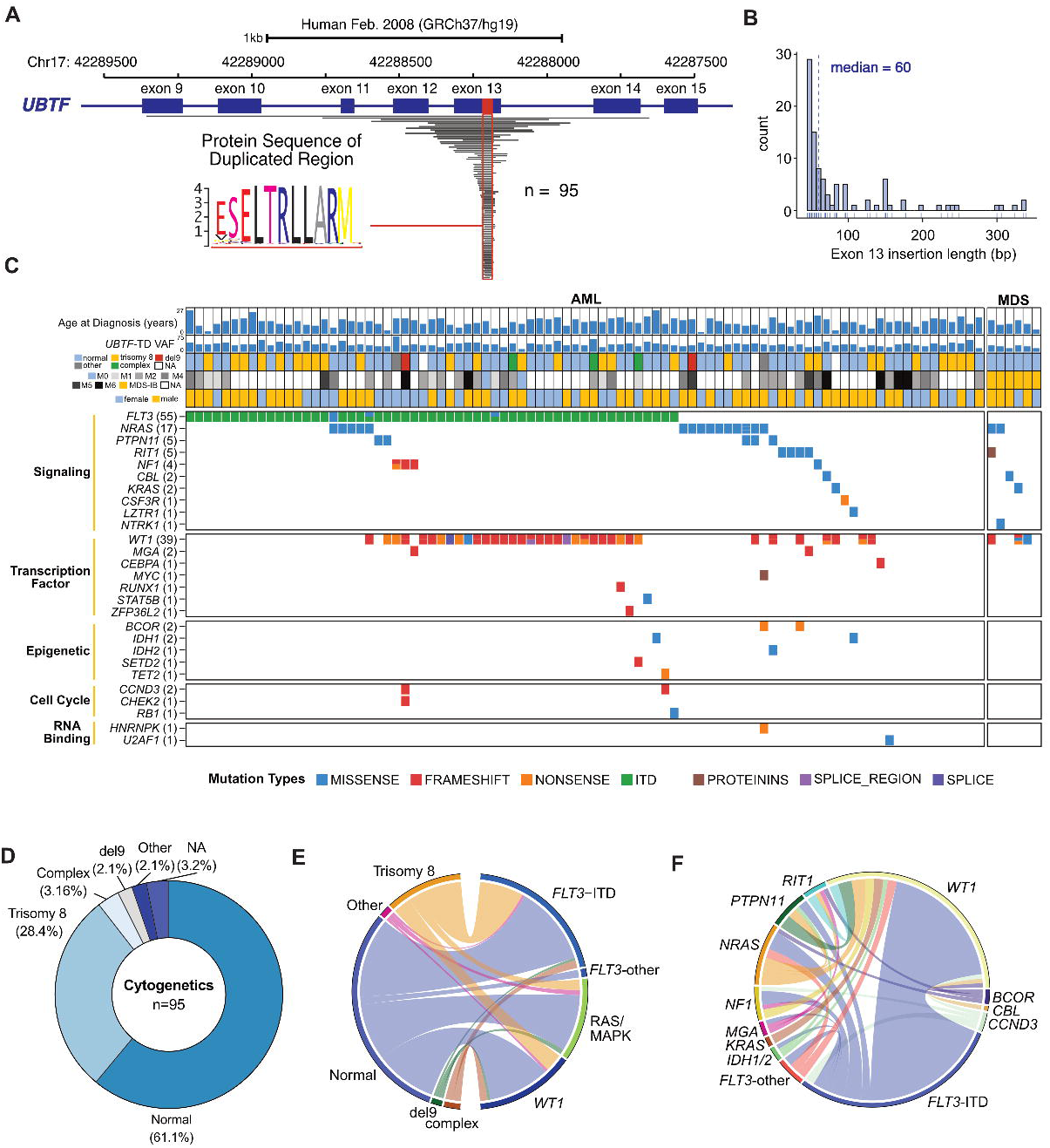
Characterization of Tandem Duplications in Exon 13 of *UBTF*. **A**. Genomic representation of duplicated regions (grey) from 95 cases with *UBTF* exon 13 duplications. A common duplicated region is outlined in red with the alignment of the resulting amino acid sequence. **B**. Size assessment of exon 13 resulting from the duplicated regions and associated insertion/deletions. For partial tandem duplications (n=13) extending beyond exon 13, sizes of duplicated exon 13 with insertions or deletions are included. **C**. Genomic landscape of *UBTF-*TD cases. For each case, variant allele frequency (VAF), diagnosis, karyotype, and sex are shown. **D**. Distribution of cytogenetic changes among *UBTF-*TD cases. **E**. Ribbon plot depicting association of cytogenetics with mutations common in *UBTF-*TD myeloid neoplasms. Genes coding for proteins involved in RAS/MAPK pathway are grouped together (*NF1, PTPN11, NRAS, KRAS, RIT1, CBL)*. All other mutations in *FLT3* not classified as ITD were grouped into *FLT3-*other. **F.** Ribbon plot depicting the association of co-occurring mutations in the *UBTF-*TD cohort. Genes with only one occurrence were excluded.

To address clonal evolution in *UBTF-*TD myeloid neoplasms, we utilized a droplet-based single cell multi-omics platform from MissionBio (**Figure 2**)(26). This platform enables the concurrent detection of UBTF-TD alterations and somatic mutations present by a custom targeted DNA panel and cell classification by using DNA-oligo conjugated antibodies targeting cell surface markers at the single cell level (**Supplemental Table 2**). In a single timepoint AML case, we found a clonal *UBTF-*TD alteration in the myeloid population and subclonal mutations in *NRAS* (*p*.G12D), *FLT3* (*p*.V592D), and *WT1* (*p*.359fs), suggesting *UBTF*-TD as an initiating event (**Supplemental Table 3, Figure 2A-B**). A *WT1* mutation was broadly present in the myeloid population, while *NRAS* or *FLT3* mutations were subclonal to *WT1* in a mutually exclusive manner, representing branched evolution of the UBTF-TD-*WT1*+ clone. We also found that cells with *WT1*^+^*FLT3*^+^ mutations were associated with high stem cell marker expression (CD34, CD117, or CD123) compared with the *UBTF*-TD-only population, whereas the *WT1*^+^*NRAS*^+^ population was characterized by low expression of these markers. In a diagnosis and relapse-paired case, we found that the identical *UBTF-*TD was retained through disease progression along with a *FLT3*-ITD alteration (**Figure 2C-D**). Interestingly, a minor *WT1*^+^ (*p.*R375fs) subclone at diagnosis was eradicated after chemotherapy, whereas a different *WT1*^+^ (*p.*R353fs) clone became dominant at relapse, showing high expression of CD34, CD117, and CD123. These data collectively confirm that *UBTF-*TD is an early initiating event, while somatic mutations are subclonal to *UBTF*-TD, possibly contributing to disease progress toward subclones with unique expression profiles.

**Figure 2.**
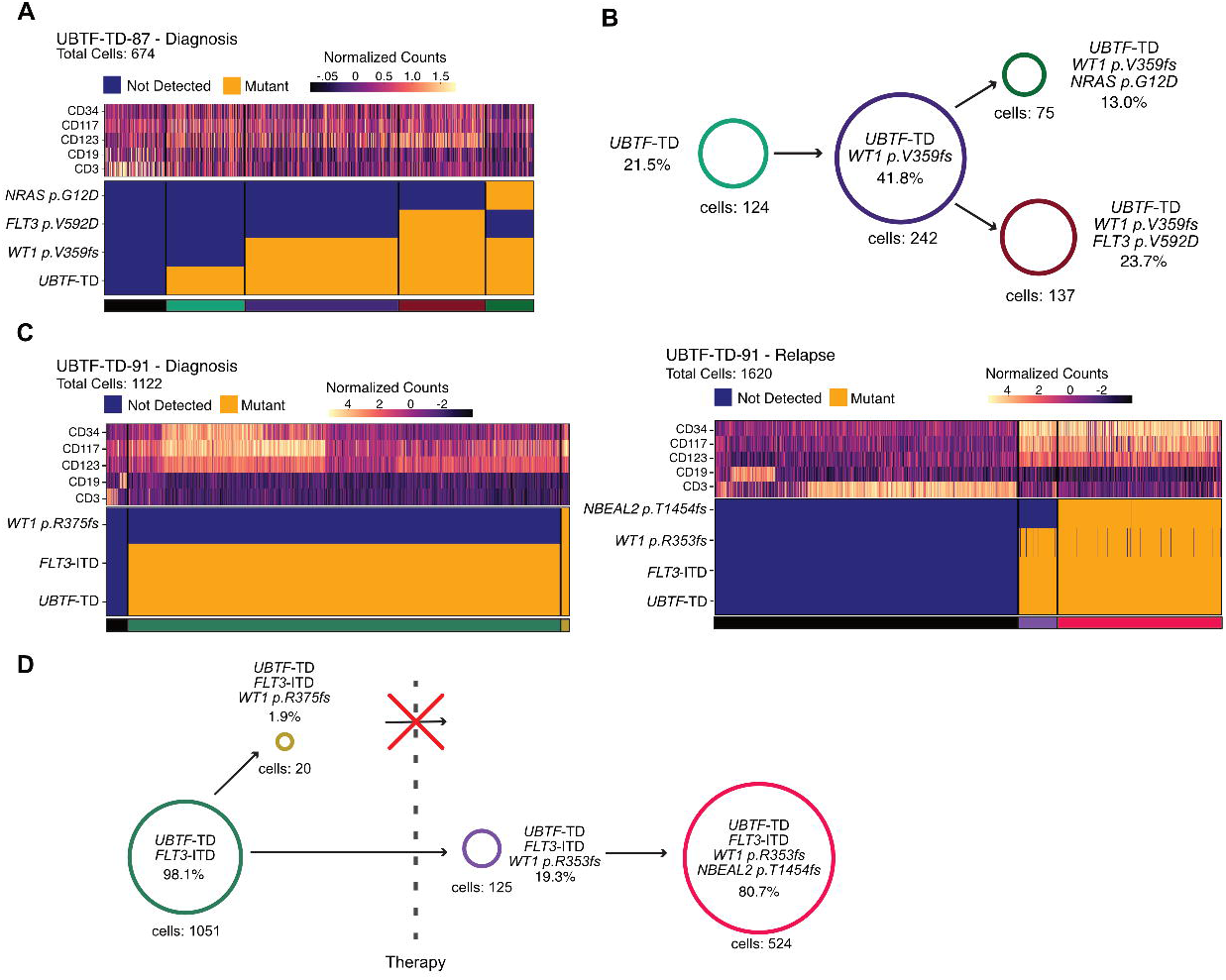
Clonal Dynamics of *UBTF-*TD Leukemias. **A.** Single-cell DNA sequencing coupled with surface marker expression of *UBTF-*TD case at diagnosis. Heatmap depicting the presence of mutant allele (bottom) and relative protein abundance (top). **B.** Schematic of clonal structure in *UBTF-*TD case from A. Percentages are calculated as a proportion of total cells with a somatic mutation. **C.** Single-cell DNA sequencing of a *UBTF-*TD case with diagnosis (left) and relapse (right) paired samples. Heatmap depicts the presence of a mutant allele and protein expression. **D.** Schematic of clonal dynamics in diagnosis/relapse case from C. Percentages are calculated as a proportion of total cells with a somatic mutation.

### Clinical features of UBTF-TD pediatric myeloid neoplasms

*UBTF*-TD AML showed a variety of morphologic features associated with cellular differentiation, as evidenced by variable FAB (French-American-British) classifications (**Figure 3A, Supplemental Table 4**). Although AML with maturation (FAB M2) was the most common (19/43, 44.2%), cases with FAB M6 (erythroid) features were also observed, as was also recently described for *UBTF*-TD AMLs in adults(9). Morphologic assessments also revealed that *UBTF-*TD cases often displayed pleomorphic blasts (**Figure 3B**), accompanied by background multilineage dysplasia and increased erythroid precursors. *UBTF*-TD AMLs showed lower white blood cell count (median = 8.4×10^9^/L, IQR = 41.4×10^9^/L) and bone marrow blast percentage (mean = 33%, IQR = 41.4%) when compared to other AMLs, including those with similar transcriptional profiles like AML with *NUP98*-rearrangements or *NPM1* mutations(1, 4) (**Figure 3C-D**). *FLT3*-ITD, but not *WT1* mutations, were associated with a higher WBC count and bone marrow blasts in *UBTF*-TD AMLs (**Figure 3E**). Despite the presence of dysplastic features, cytogenetic studies commonly found either a normal karyotype (58/95, 61.1%) or trisomy 8 (27/95, 28.4%), and myelodysplasia-related chromosomal changes or myelodysplasia-related mutations were overall rare, suggesting that *UBTF*-TD itself contributes to dysplastic features (**Supplemental Table 4**). Considering these overall features, the majority of UBTF-TD AMLs (83/89, 93.3%) are best classified as “Acute myeloid leukemia with other defined genetic alterations” in the current WHO classification(25) (**Supplemental Table 4**).

**Figure 3.**
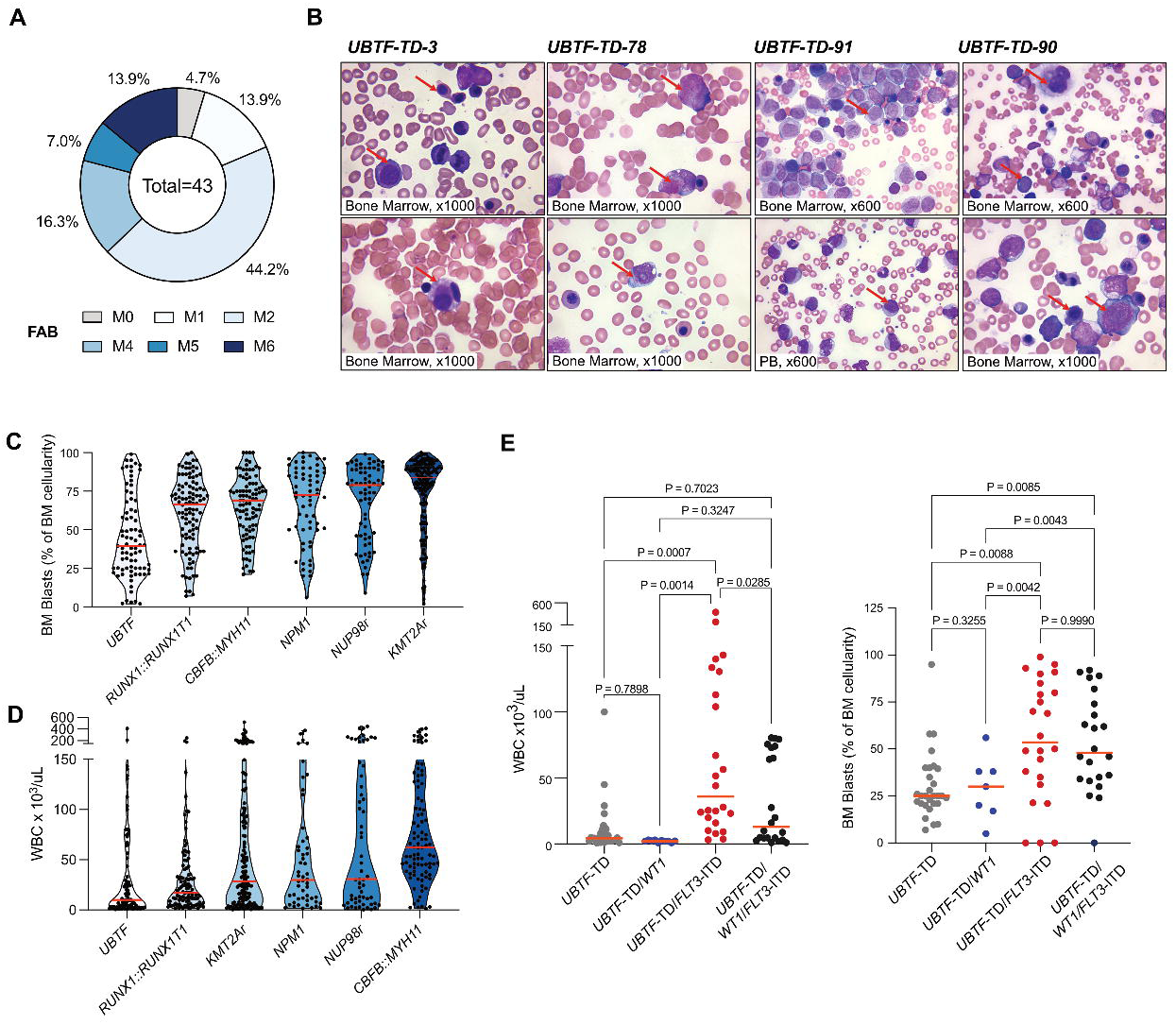
Morphological and Biological Assessment of *UBTF-*TD. **A**. Available FAB (French-American-British) classification of *UBTF-*TD AMLs (n=43). **B**. Representative Wright-Giemsa Staining of bone marrow (BM) aspirates and peripheral blood (PB) smears from 4 unique *UBTF-*TD cases. Case ID, FAB/WHO classification, along with *FLT3* and *WT1* mutation status, are labeled above. UBTF-TD-3-**Childhood MDS with increased blasts**: increased erythroid cells with dysplastic morphologic features (top), dysplastic myeloid cells with salmon-colored granules, and dysplastic small megakaryocytes (bottom). **UBTF-TD-78**-**AML with other defined genetic alterations (Acute myeloid leukemia with maturation)**: characteristic blasts and myeloid precursor cells with salmon-colored granules (top) and blasts with small Auer rods (bottom). **UBTF-TD-91-AML with other defined genetic alterations (Acute myelomonocytic leukemia)**: blasts with both myeloid and monocytic/monoblastic morphologic features. PB in this patient is one of the few cases in our cohort showing hyperleukocytosis with many circulating blasts (bottom). **UBTF-TD-90-AML with other defined genetic alterations (Acute myelomonocytic leukemia)**: dysplastic megakaryocytes, increased erythroid cells with dysplastic morphologic features, and blasts, many with monocytic/monoblastic morphologic features. **C.** White blood cell counts (WBC) for a pediatric AML cohort stratified by oncogenic driver subtypes. **D.** Bone marrow blast percentage for a pediatric AML cohort stratified by oncogenic driver subtypes. **E.** WBC count and BM blast percentage among *UBTF*-TD cases with different *WT1* and *FLT3* mutation status.

### Transcriptional features of UBTF-TD myeloid neoplasms

We and others have previously shown that AML with *UBTF*-TD is characterized by high *HOXA* and *HOXB* cluster gene expression, similar to *NPM1*-mutated or *NUP98*::*NSD1* AML (1, 4, 8). To further define the unique expression profiles of *UBTF-*TD, we established an RNA-seq cohort consisting of various AML subtypes (1) (n=837), cord blood CD34+ samples from healthy donors (n=5), and *UBTF-*TD AML and MDS samples (n=94: 1 *UBTF-*TD case did not have RNA-seq data available) (**Figure 4A**). Consistent with previous data, *UBTF-*TD cases clustered with *NPM1-*mutated and *NUP98::NSD1* AMLs. However, *UBTF-*TD displayed a significantly higher expression of a subset of *HOXB* cluster genes (e.g., *HOXB8, HOXB9*) compared with *NUP98*::*NSD1* AML (**Figure 4B**). We also observed uniquely high expression of histone genes (e.g., *HIST1H4F* and *HIST1H2B1*) compared to *NPM1*-mutated AML (**Figure 4C**), suggesting transcriptional mechanisms unique to *UBTF*-TD AML. Within *UBTF-*TD samples, those with *FLT3*-ITD and *WT1* mutations showed unique distribution on the UMAP cluster (**Figure 4D**), and each mutation group demonstrated differential gene expression against *UBTF*-TD samples without either mutation (**Figure 4E**). Co-occurrence of *WT1* and *FLT3*-ITD was associated with stemness-related genes (e.g., *CD34* and *DNTT,* **Figure 4E-F**), and Gene Set Enrichment Analysis (GSEA) confirmed enrichment of stemness or cell cycle-related gene expression in *WT1*^+^*FLT3*-ITD^+^ *UBTF*-TD samples (**Figure 4G**). These results show the unique expression profile of *UBTF*-TD AML and the specific influence of additional cooperating mutations, which can likely influence patterns of clonal evolution.

**Figure 4.**
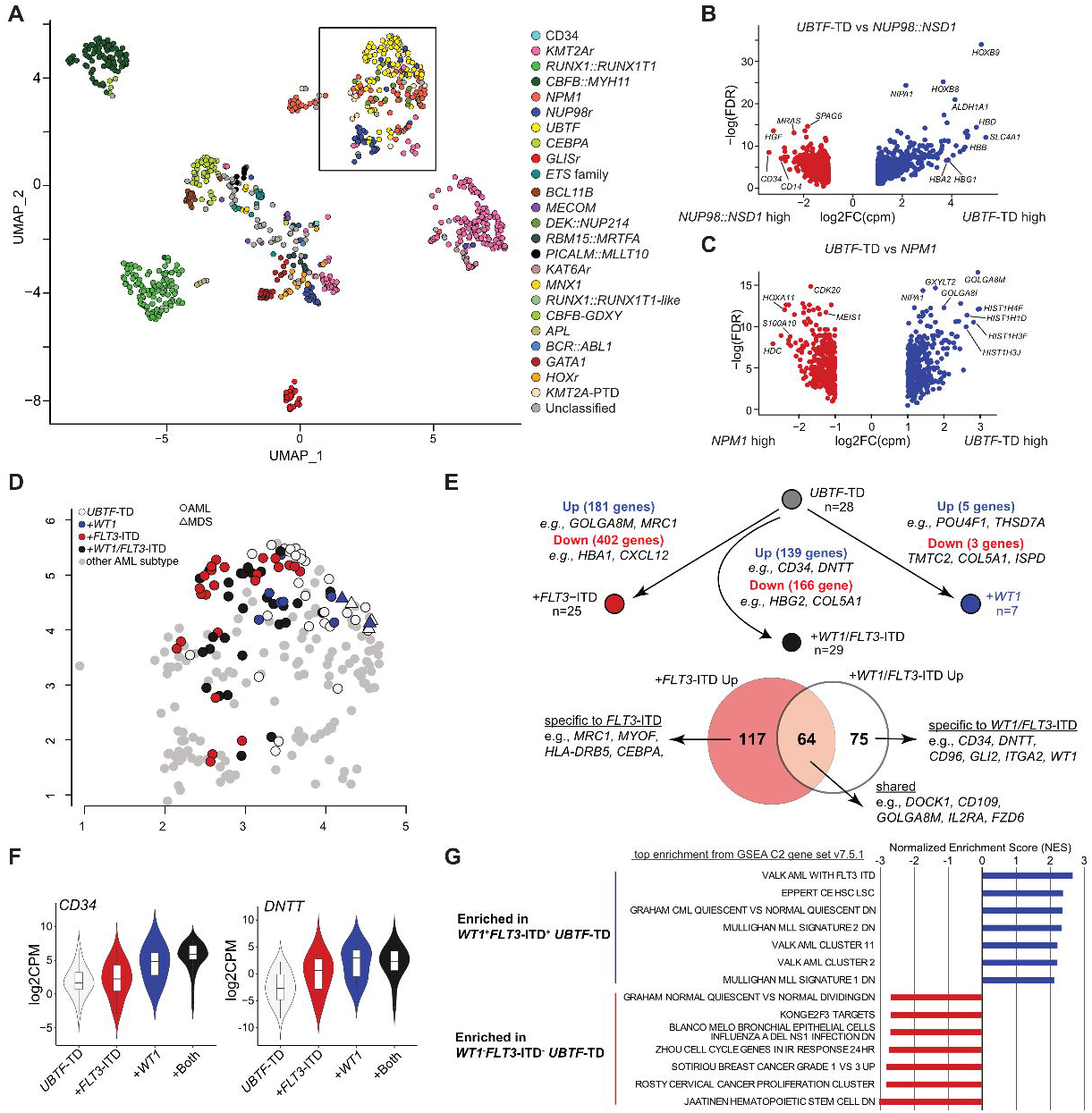
Transcriptional characterization of *UBTF-*TD leukemias. **A**. UMAP (Uniform Manifold Approximation and Projection) of expression profiles across a cohort of *UBTF*-TD (n=94) and pediatric AML (n=839), adapted from a previous study(4). Each dot is colored by subtype-defining alterations. The black box outlines a cluster of cases with *HOXA/HOXB* dysregulation, which includes all *UBTF*-TD cases. **B.** Differentially expressed genes between *UBTF*-TD AML and *NUP98::NSD1* pediatric AML. Representative genes are annotated. **C**. Differentially expressed genes between *UBTF*-TD AML and *NPM1-*mutated pediatric AML. **D.** Distribution of *UBTF*-TD cases within the cluster with HOXA/B dysregulation. Each data is colored by the mutational status of *UBTF-*TD, *FLT3-*ITD, or *WT1*. MDS cases are depicted with triangles, and AML cases are with circles. **E-top.** Schematic depicting the transcriptional comparison of *UBTF-*TD cases by mutational status. **-bottom.** Venn diagram showing overlap of differentially expressed genes in *FLT3*-ITD only cases and *FLT3*-ITD+/*WT1*+ cases compared with cases without these mutations. **F.** Expression *CD34* and *DNTT* with respect to *FLT3* and *WT1* mutational status. **G.** GSEA of *UBTF*-TD cases based on mutational status.

### Exon 9 tandem duplications in UBTF

Given the recurrent *UBTF* exon 13 alterations duplicating specific hydrophobic amino acid sequences, we hypothesized that *UBTF* alterations outside exon 13 resulting in similar amino acid sequences could be found in cases without defining alterations but with a similar expression signature. By close inspection of the *UBTF* gene using RNA-sequencing data, we found two pediatric AML cases without exon 13 *UBTF*-TD or other driver alterations that instead have in-frame tandem duplications (lengths of 78 and 153bp) in exon 9 of *UBTF* (TD-exon9), encoding short hydrophobic amino acid sequences (**Figure 5A-B**). These cases express *HOXA/B* cluster genes comparably to exon 13 *UBTF*-TD (**Supplemental Figure 1**), and one had a *WT1* mutation (**Supplemental Table 5**). To test whether these exon 9 duplication events could lead to leukemic transformation, we expressed both exon 9 duplications in cbCD34+ cells using lentiviral vectors and assessed their impact on cell proliferation, clonogenic potential, and cellular morphology in comparison with control conditions and exon 13 UBTF-TD (**Figure 5C-D**). Colony forming unit (CFU) assay revealed that expression of both UBTF-TD-exon9 increased total colony number (**Figure 5E**). After the second round of replating, cells with UBTF-TD-exon9 showed an immature morphology along with erythroid features, similar to exon 13 UBTF-TD expressing cells (**Figure 5F**). Furthermore, cells expressing UBTF*-*TD-exon9 as well as UBTF-TD-exon13 showed a proliferative advantage compared to UBTF-WT and vector controls (**Figure 5G**). Collectively, these data highlight a tandem duplication in *UBTF* exon 9 as a defining alteration functionally equivalent to exon 13 tandem duplications.

**Figure 5.**
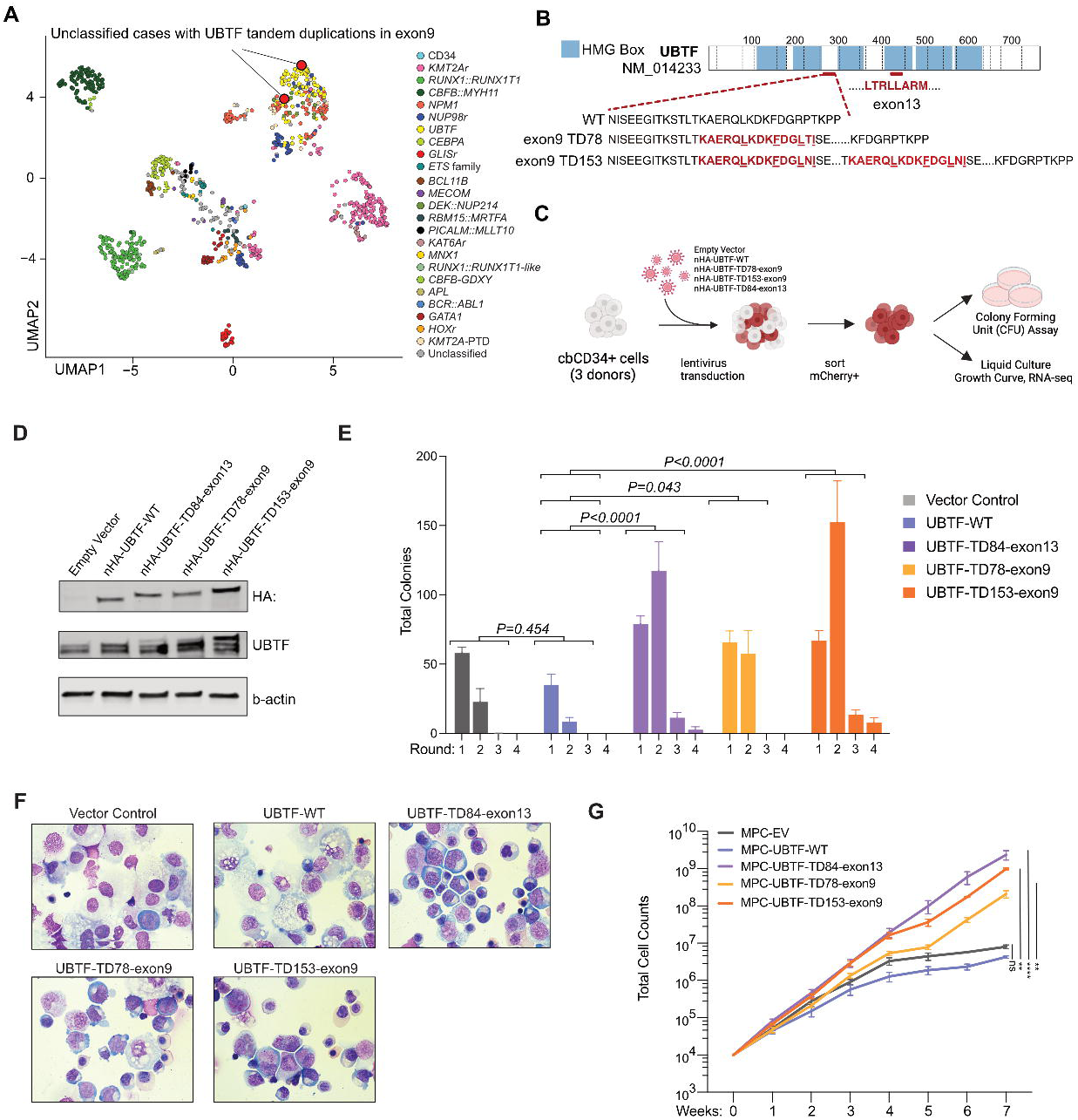
Exon 9 *UBTF* Tandem Duplications in Pediatric AML. **A.** Identification of *UBTF-*TD cases with tandem duplications in exon 9 of *UBTF. UBTF-*TD-exon9 on the UMAP plot of the Pediatric AML cohort (red circles), adapted from a previous study(4). **B.** Schematic of UBTF protein, highlighting amino acid sequences encoded in exon 9 of *UBTF*-WT, *UBTF-*TD78-exon9, *UBTF-*TD153-exon9. Duplications resulting in short hydrophobic sequences are labeled in red. Hydrophobic residues are underlined. **C.** Experimental design to evaluate the transforming potential of UBTF*-*TD-exon9 mutants **D.** Immunoblot of lysates from cbCD34+ transduced with UBTF-TD-exon9 vectors and corresponding controls. Antibodies against HA-tag, UBTF, and β-actin were used. **E**. Colony forming unit assay. 2-way ANOVA with two-sided Dunnett’s test was applied with UBTF-WT as control **F.** Cytospins isolated from CFU assay at round 2 of replating (Wright-Giemsa Staining, 400X magnification). **G.** In vitro growth curves of cbCD34+ cells expressing controls or UBTF*-*TD-e9 mutants. 2-way ANOVA with Dunnett’s test was applied with UBTF-WT as control.

## Discussion

In this study, we extended our cohort of pediatric myeloid malignancies with exon 13 *UBTF*-TD, which now includes 95 pediatric cases. Similar to studies in adults (9), pediatric myeloid tumors with *UBTF-*TD are associated with lower bone marrow blast infiltration and lower white blood cell count, suggesting a similar clinical presentation. In this pediatric cohort, we observed that *UBTF-*TD is associated with an M2 morphology subtype (44.2%) according to FAB classification (27). These morphological features align with findings in adult cases, which showed a high prevalence of FAB M6 and M2 cases (9). Furthermore, the presence of dysplastic features and the observation that *UBTF*-TD occurs in cases of pediatric MDS suggest that MDS and AML could be part of a continuum driven by *UBTF*-TD. Progression to AML may require the acquisition of cooperating mutations such as *FLT3-*ITD, supported by the finding that none of the MDS cases in this cohort harbor a *FLT3*-ITD alteration.

To evaluate the impact of these cooperating mutations on cellular status, we analyzed RNA-seq data of *UBTF*-TD cases with or without mutations, showing that the co-occurrence of *FLT3-*ITD and *WT1* is strongly associated with stem cell features and represented by CD34 expression or quiescent states. These findings were further supported by single cell studies. Given that *WT1* mutations are often subclonal alterations that expand at relapse and that the co-occurrence of these mutations is associated with particularly dismal outcomes (4), acquiring these stem-like features can contribute to resistance to conventional chemotherapy. Furthermore, a recent study showed that UBTF-TD localizes to HOXA/B genomic regions and that Menin inhibition is sufficient to promote terminal differentiation (28). How these other mutations co-operate with UBTF-TD to maintain stem cell features requires further evaluation.

Given that exon 13 *UBTF*-TD has been underappreciated in standard computational pipelines, we investigated other possible *UBTF* alterations in pediatric AML cases without a defined driver event and demonstrated that in-frame tandem duplications in exon 9 of *UBTF* are possible driver alterations. Although rare within cases with *UBTF* alterations (2/97 *UBTF-*TD cases, 2.1%), cases with tandem duplications in exon 9 show similar transcriptional profiles to exon 13 *UBTF*-TD AML. We further show that exon 9 alterations can induce leukemic changes, including hematopoietic cell growth and increased clonogenicity in cbCD34+ cells similar to exon 13 alterations. At the amino acid level, exon 9 tandem duplications contain amino acid sequences (LKDKFDGLTI) that share hydrophobic α-helix structure with recurrently duplicated sequences in exon 13 tandem duplications (LTRLLARM), suggesting a shared mechanism. Importantly, PCR-based screening can underestimate PTD or these exon 9 alterations since they are outside the covered regions(9), and we propose that enhanced sequencing-based strategies are needed to accurately diagnose this entity. The findings presented here will help build on our understanding of UBTF-TD myeloid neoplasms and further support it’s recognition as a distinct entity in future classification systems.

## Data Availability

The expression data newly generated in this study (RNA-Seq: n=3) and scDNA + protein sequencing (n=3) have been deposited in the European Genome-Phenome Archive (EGA) which is hosted by the European Bioinformatics Institute (EBI), under accession EGAS00001005760. The remaining RNA-Seq data are available via EGA, St. Jude Cloud or TARGET as defined in Supplemental Table 6. Information about TARGET can be found at http://ocg.cancer.gov/programs/target. Other data generated in this study are available in the Supplemental tables or upon request to the corresponding author.

## Acknowledgements

The work was funded by the American Lebanese and Syrian Associated Charities of St. Jude Children’s Research Hospital and funds from the US NIH, including F32 HL154636 (JMB), U54 CA243124 and R01 CA276079 (JMK). The content, however, does not necessarily represent the official views of the NIH and is solely the responsibility of the authors. The studies were also funded by the Jane Coffin Childs Fund (JMB). JMK holds a Career Award for Medical Scientists from the Burroughs Welcome Fund. Support was also provided by Shared Resources provided through the St. Jude Comprehensive Cancer Center (P30-CA21765), Flow Cytometry and Cell Sorting, Comparative Pathology Core, and Genome Sequencing (Hartwell Center).

## Author Contributions

Contributions: Conceptualization, J.M.K; Methodology, J.M.B., L.C., E.X.; Software, Validation, J.M.B., L.C., E.X.; Formal Analysis, J.M.B., M.U., B.O., M.A.L., M.K.; Investigation, J.M.B., M.U., L.C., M.P.W., M.K., M.E.T., X.M.; Resources, J.M.K.; Data Curation, T.W., G.S.; Writing – Original Draft, J.M.B., M.U.; Writing – Review & Editing, all authors were involved in review & editing of the manuscript; Visualization, J.M.B., M.U.; Supervision, J.M.K., Project Administration, J.M.B., M.U., T.W., S.A., J.M.K.; Funding Acquisition, J.M.B., J.M.K.

## Competing Interests

C.Y., B.O., and M.A. are employed by Mission Bio, Inc. No other authors have conflicts to declare.

## Data Availability Statement

Samples from patients with MDS or AML from St. Jude Children’s Research Hospital tissue resource core facility were obtained with written informed consent using a protocol approved by the St. Jude Children’s Research Hospital institutional review board (IRB). Studies were conducted in accordance with the International Ethical Guidelines for Biomedical Research Involving Human Subjects. The expression data newly generated in this study (RNA-Seq: n=3) and scDNA + protein sequencing (n=3) have been deposited in the European Genome-Phenome Archive (EGA) which is hosted by the European Bioinformatics Institute (EBI), under accession <u>EGAS00001005760</u>. The remaining RNA-Seq data are available via EGA, St. Jude Cloud or TARGET as defined in **Supplemental Table 6**. Information about TARGET can be found at http://ocg.cancer.gov/programs/target. Other data generated in this study are available in the Supplemental tables or upon request to the corresponding author.

## Notes

### Author Declarations

Samples from patients with MDS or AML from St. Jude Childrens Research Hospital tissue resource core facility were obtained with written informed consent using a protocol approved by the St. Jude Childrens Research Hospital institutional review board (IRB). Studies were conducted in accordance with the International Ethical Guidelines for Biomedical Research Involving Human Subjects

## Citations

1. Umeda M, Ma J, Westover T, Ni Y, Song G, Maciaszek J, et al. Proposal of a new genomic framework for categorization of pediatric acute myeloid leukemia associated with prognosis. Res Sq. 2023.

2. Schwartz JR, Ma J, Lamprecht T, Walsh M, Wang S, Bryant V, et al. The genomic landscape of pediatric myelodysplastic syndromes. Nat Commun. 2017;8(1):1557.

3. Bolouri H, Farrar JE, Triche T, Jr., Ries RE, Lim EL, Alonzo TA, et al. The molecular landscape of pediatric acute myeloid leukemia reveals recurrent structural alterations and age-specific mutational interactions. Nat Med. 2018;24(1):103–12.

4. Umeda M, Ma J, Huang BJ, Hagiwara K, Westover T, Abdelhamed S, et al. Integrated genomic analysis identifies UBTF tandem duplications as a recurrent lesion in pediatric acute myeloid leukemia. Blood Cancer Discov. 2022.

5. Kaburagi T, Shiba N, Yamato G, Yoshida K, Tabuchi K, Ohki K, et al. UBTF-Internal tandem duplication as a novel poor prognostic factor in pediatric acute myeloid leukemia. Genes Chromosomes Cancer. 2022.

6. Stratmann S, Yones SA, Mayrhofer M, Norgren N, Skaftason A, Sun J, et al. Genomic characterization of relapsed acute myeloid leukemia reveals novel putative therapeutic targets. Blood Adv. 2021;5(3):900–12.

7. Ma X, Liu Y, Liu Y, Alexandrov LB, Edmonson MN, Gawad C, et al. Pan-cancer genome and transcriptome analyses of 1,699 paediatric leukaemias and solid tumours. Nature. 2018;555(7696):371–6.

8. Georgi JA, Stasik S, Eckardt JN, Zukunft S, Hartwig M, Rollig C, et al. UBTF tandem duplications are rare but recurrent alterations in adult AML and associated with younger age, myelodysplasia, and inferior outcome. Blood Cancer J. 2023;13(1):88.

9. Duployez N, Vasseur L, Kim R, Largeaud L, Passet M, L’Haridon A, et al. UBTF tandem duplications define a distinct subtype of adult de novo acute myeloid leukemia. Leukemia. 2023.

10. Sanij E, Hannan RD. The role of UBF in regulating the structure and dynamics of transcriptionally active rDNA chromatin. Epigenetics. 2009;4(6):374–82.

11. Moss T, Mars JC, Tremblay MG, Sabourin-Felix M. The chromatin landscape of the ribosomal RNA genes in mouse and human. Chromosome Res. 2019;27(1-2):31–40.

12. Li H, Durbin R. Fast and accurate long-read alignment with Burrows-Wheeler transform. Bioinformatics. 2010;26(5):589–95.

13. Li H, Handsaker B, Wysoker A, Fennell T, Ruan J, Homer N, et al. The Sequence Alignment/Map format and SAMtools. Bioinformatics. 2009;25(16):2078–9.

14. Wu G, Diaz AK, Paugh BS, Rankin SL, Ju B, Li Y, et al. The genomic landscape of diffuse intrinsic pontine glioma and pediatric non-brainstem high-grade glioma. Nat Genet. 2014;46(5):444–50.

15. Tian L, Li Y, Edmonson MN, Zhou X, Newman S, McLeod C, et al. CICERO: a versatile method for detecting complex and diverse driver fusions using cancer RNA sequencing data. Genome Biol. 2020;21(1):126.

16. Edmonson MN, Zhang J, Yan C, Finney RP, Meerzaman DM, Buetow KH. Bambino: a variant detector and alignment viewer for next-generation sequencing data in the SAM/BAM format. Bioinformatics. 2011;27(6):865–6.

17. Hagiwara K, Edmonson MN, Wheeler DA, Zhang J. indelPost: harmonizing ambiguities in simple and complex indel alignments. Bioinformatics. 2022;38(2):549–51.

18. Hagiwara K, Ding L, Edmonson MN, Rice SV, Newman S, Easton J, et al. RNAIndel: discovering somatic coding indels from tumor RNA-Seq data. Bioinformatics. 2020;36(5):1382–90.

19. Anders S, Pyl PT, Huber W. HTSeq--a Python framework to work with high-throughput sequencing data. Bioinformatics. 2015;31(2):166–9.

20. Ritchie ME, Phipson B, Wu D, Hu Y, Law CW, Shi W, et al. limma powers differential expression analyses for RNA-sequencing and microarray studies. Nucleic Acids Res. 2015;43(7):e47.

21. Satija R, Farrell JA, Gennert D, Schier AF, Regev A. Spatial reconstruction of single-cell gene expression data. Nat Biotechnol. 2015;33(5):495–502.

22. Becht E, McInnes L, Healy J, Dutertre CA, Kwok IWH, Ng LG, et al. Dimensionality reduction for visualizing single-cell data using UMAP. Nat Biotechnol. 2018.

23. Subramanian A, Tamayo P, Mootha VK, Mukherjee S, Ebert BL, Gillette MA, et al. Gene set enrichment analysis: a knowledge-based approach for interpreting genome-wide expression profiles. Proc Natl Acad Sci U S A. 2005;102(43):15545–50.

24. Fornerod M, Ma J, Noort S, Liu Y, Walsh MP, Shi L, et al. Integrative Genomic Analysis of Pediatric Myeloid-Related Acute Leukemias Identifies Novel Subtypes and Prognostic Indicators. Blood Cancer Discov. 2021;2(6):586–99.

25. Khoury JD, Solary E, Abla O, Akkari Y, Alaggio R, Apperley JF, et al. The 5th edition of the World Health Organization Classification of Haematolymphoid Tumours: Myeloid and Histiocytic/Dendritic Neoplasms. Leukemia. 2022;36(7):1703–19.

26. Miles LA, Bowman RL, Merlinsky TR, Csete IS, Ooi AT, Durruthy-Durruthy R, et al. Single-cell mutation analysis of clonal evolution in myeloid malignancies. Nature. 2020;587(7834):477–82.

27. Bennett JM, Catovsky D, Daniel MT, Flandrin G, Galton DA, Gralnick HR, et al. Proposed revised criteria for the classification of acute myeloid leukemia. A report of the French-American-British Cooperative Group. Ann Intern Med. 1985;103(4):620–5.

28. Barajas JM, Rasouli M, Umeda M, Hiltenbrand RL, Abdelhamed S, Mohnani R, et al. Acute myeloid leukemias with UBTF tandem duplications are sensitive to Menin inhibitors. Blood. 2023.

