## Supplemental Figures for "*UBTF* Tandem Duplications in Pediatric MDS and AML: Implications for Clinical Screening and Diagnosis"

**Supplemental Figure 1.** *UBTF-*TD exon9 AMLs have a similar expression profile to exon13 *UBTF-*TD AMLs. **A.** Scatterplot comparing gene expressing of *UBTF-*TD exon9 AMLs to UBTF-TD exon13 AMLs. **B.** Gene expression comparison of *UBTF-*TD exon9 AMLs to other AML subtypes. **C.** Gene expression comparison of *UBTF-*TD exon13 AMLs to other AML subtypes. **D.** Heatmap depicting *HOXA/HOXB* gene expression across AML molecular categories.
